# Association Between Diabetes Mellitus and Endodontic Pathosis

**DOI:** 10.1101/2023.08.16.23294173

**Authors:** Kortnie K. Strother, Krishna Kumar Kookal, Vashti Bueso, Muhammad F. Walji, Ariadne Letra

**Author notes:** **Corresponding Author (current address) Ariadne Letra, DDS, MS, PhD**, Professor, Department of Oral and Craniofacial Sciences, Center for Craniofacial and Dental Genetics, Bridgeside Point I, 100 Technology Drive, room 450L, Pittsburgh PA, 15219. **Authors’ contributions:** KK contributed to data acquisition and analysis, drafted the manuscript, VS, KKK, MFW contributed to data acquisition, critically reviewed the manuscript, AL contributed to study conception and design, drafted the manuscript. All authors gave final approval of the version to be submitted.

## Abstract

**Background:** Studies have shown that periodontal and periapical diseases are more prevalent among diabetes mellitus (DM) patients. Here, we evaluated the potential relationship between endodontic pathologies and DM through a retrospective analysis of 2,000 electronic health records (EHR) of individuals with/without DM.

**Methods:** Records of patients treated at UTHealth School of Dentistry presenting with a history of endodontic treatment with and without DM were randomly selected for analysis of 24 treatment- and patient-centric variables to assess for the association between endodontic disease and DM. Data between groups were compared using Chi-square, Fisher Exact tests, and one-way analysis of variance (ANOVA). Significant differences were set at P<0.05.

**Results:** Diabetic patients had significantly less symptomatic pulpal diagnoses, particularly less symptomatic irreversible pulpitis (P<0.00005); whereas they had significantly more symptomatic apical periodontitis (P=0.01) than non-diabetics. Diabetics had a significantly greater number of endodontically-treated teeth (P=2.2 × 10^−16^), particularly canines and molars (P<0.002). The frequency of active periapical lesions was higher in non-diabetics, although no differences in lesion size were observed. The frequency of periodontal disease and additional systemic disease(s), and smoking was significantly increased in DM patients with endodontic treatment (P<0.05). No differences were observed between the number of medications taken between groups or upon stratifying analysis by gender (P>0.05).

**Conclusions:** Our results continue to support that DM contributes to increased endodontic disease and highlight the complex relationships between oral and systemic diseases.

**Practical Implications:** Dental professionals should be cognizant of endodontic pathology as a clinical complication of DM that presents as more chronic in nature, at a greater frequency, and have a tendency toward a nonhealing outcome.

## INTRODUCTION

Diabetes mellitus (DM) is a common metabolic disorder affecting more than 30 million individuals in the United States.^1^ DM is characterized by deficient or absent insulin production by the pancreas leading to dysfunctional metabolism of carbohydrates, protein or fat.^2–4^ Type I DM comprises about 5% of individuals with DM and is an autoimmune reaction that destroys pancreatic β-cells, thus resulting in absolute insulin insufficiency. Type II DM accounts for 90%-95% of individuals with DM and is characterized by β-cell dysfunction and tissue resistance to insulin action. Other forms of DM involving the body’s ability to produce, secrete, and/or response to insulin include gestational diabetes, monogenic diabetes syndromes, cystic fibrosis-related diabetes, and post-transplantation DM.^1–3,5^

Individuals with DM (particularly those with poorly controlled disease) are known to bear systemic complications more often than those without. DM triggers a state of chronic inflammation in which advanced glycation end-products and pro-inflammatory cytokines are elevated while macrophage growth factors are decreased. Together, these alterations impair leukocyte function while increased the inflammatory response to contribute to poor wound healing and increased susceptibility to infection.^5–9^ Alterations in bone metabolism, peripheral neuropathy, vascular insufficiency and autonomic dysfunction, which in turn may lead to dysfunction of the nerves, blood vessels and wound healing capabilities, have been frequently described in individuals with DM.^2^ Further, DM patients are also more prone to oral infections and complications such as caries, hyposalivation, taste changes, oral candidiasis, halitosis and more.^2,6,7,10–12^

Although an association between periodontitis and DM is well-established, the relationship between DM and endodontic pathosis (particularly apical periodontitis, AP) is not as clear, and controversial findings have been described.^2,6,11,12^ AP develops in the periradicular tissue as a result of pulp necrosis and the presence of endodontic pathogens within the root canal system with subsequent release of endotoxins through the apical foramen activate the host innate immune response to increase cytokine production and trigger osteoclastogenesis leading to bone resorption.^13–15^ Previous studies have assessed the relationship between DM and AP using a variety of approaches including direct comparisons of the prevalence of AP, levels of glycated hemoglobin levels (HbA1c), or by evaluating cytokine expression levels in AP tissues of diabetic and non-diabetic patients.^2,7,9,10,16–18^ Overall, a higher prevalence of AP has been reported in diabetics than in non-diabetics (74% versus 42%, respectively)^19^, and periapical status seemed to be significantly associated with HbA1c > 6.5%^20^, although findings are not consistent across the studies. Differences in the populations studied, study design and variables analyzed, may have accounted for such differences which often make direct comparison among the studies difficult. Recently, two systematic reviews provided additional evidence supporting a positive relationship between AP and DM, and concluded that DM patients showing a 3-fold greater chance of having AP than those without DM in both untreated and endodontically-treated teeth.^2,7,9,10,16–18^ Increased expression of proinflammatory cytokines such as IL-1α, IL-1β, IL-2, IL-6, IL-12, TNF-α, IFN-γ, and CRP in AP tissues of diabetics compared to nondiabetics has also been reported.^14,15,21^ Despite these observations, the relationship between endodontic disease and DM remains inconclusive.

In this study, we performed a retrospective analysis of 2,000 patient electronic health records (EHRs) to further investigate the potential association between DM and endodontic pathosis. A cross-sectional study design was used to compare the frequencies of twenty-four patient- and treatment-centered variables in diabetic and non-diabetic patients. Our findings continue to support that DM contributes to increased predisposition to pulpal and periapical disease.

## MATERIALS AND METHODS

### Study population

This study was approved by the UTHealth Committee for Protection of Human Subjects (HSC-DB-17-0565). We randomly selected 2,000 electronic health records (aXium, Exan Software) between 01/01/2008 and 03/31/2017 to identify patients treated at the UTHealth School of Dentistry (UTSD) presenting with (n=500/group): 1) history of endodontic treatment and DM (Endo+DM+), 2) history of endodontic treatment without DM (Endo+DM-); 3) no history of endodontic treatment with DM (Endo-DM+), and 4) no history of endodontic treatment without DM (Endo-DM-). Records were included for subjects 18 years of age or older, with or without a diagnosis of endodontic pathology, and with or without an individual history of diabetes or pre-diabetes. The determination of diabetes was made using the self-reported history of DM within the medical record as a “yes” or “no” response. Further information indicating the type of diabetes was assessed from individual records when available. The treatment codes D3000 through D3999 were used to identify patients with who received endodontic treatment of at least 1 tooth at UTSD. To minimize potential confounders in the analysis, records of individuals who were edentulous, pregnant during treatment (due to potential gestational DM), or with a history of diabetes insipidus, cancer, stroke, HIV/AIDS, seizures/epilepsy, organ transplant, or genetic disease affecting bone metabolism, were not included in the study. Records of patients who had endodontic treatment performed outside of UTSD were also excluded.

Twenty-four patient- and treatment-centered variables were assessed from each record, through careful evaluation of dental/medical history forms, odontograms, radiographs, and endodontic treatment notes (when applicable) (Tables 1-3). Data were collected by 2 evaluators (one of which is an endodontist), who were calibrated using 300 sample records and maintaining an open line of communication during data collection for 2000 records. Information was recorded without any patient identifiers into an encrypted Excel worksheet. Patient-centric valuables were: age, gender, self-reported individual and family history of DM, history of additional systemic conditions, smoking, presence/absence of periodontal disease (defined as healthy periodontium, gingivitis, localized chronic periodontitis, chronic periodontitis, based on the 1999 periodontal classification scheme)^22^, type and number of medications taken. For patient-centric variables collected from the medical history form, if the response was left blank, a “no” or “none” was assumed as these are “yes” or “no” variables that a patient or provider may not complete if they deem nonsignificant. Treatment-centric variables were: presence/absence of completed endodontic treatment at UTSD, total number of endodontically treated teeth present (based on available radiographs), endodontically-treated tooth type and location (maxilla or mandible), pulpal [symptomatic irreversible pulpitis (SIP), asymptomatic irreversible pulpitis (AIP), reversible pulpitis (RP), pulp necrosis (PN), previously treated (PT), previously initiated (PI)], and periapical diagnoses [symptomatic apical periodontitis (SAP), asymptomatic apical periodontitis (AAP), chronic apical abscess (CAA), acute apical abscess (AAA), normal apical tissue (NAT))^23^ associated with respective treatment code at UTSD, presence of absence of periapical lesion on the endodontically-treated tooth, lesion size (in millimeters, as measured on the MiPACS digital radiography viewer), healing outcome of the endodontically-treated tooth (based on treatment recall appointment notes). Healing outcome was obtained from follow-up appointment notes and classified as healed, healing, diseased/failed, based on established parameters.^24^ Teeth without a detectable periapical lesion at the start of treatment and at follow-up appointments, without any symptoms were considered healed. Teeth with detectable periapical lesion at the start of treatment but showing a reduction in lesion size or resolution, and any absence of symptoms at the follow up visit were considered healing/healed. Teeth were considered diseased if a periapical lesion increased in size, or if symptoms were present at the follow up visit. If information regarding treatment-centric variables or treatment codes was not recorded in the EHR, the data was considered missing and not included in the analysis.

**Table 1.** Frequencies of treatment-centric variables in individuals with and without DM.

| Variable | Endo+/DM-<br>N (%) | Endo+/DM+<br>N (%) | P-value* | OR (95% CI)** |
| --- | --- | --- | --- | --- |
| Mean Number of Treated Teeth | 1.8 | 2.6 | **2.2x10<sup>-16</sup>** | 0.08-0.11 |
| *Pulpal Diagnosis* |  |  |  |  |
| Pulp Necrosis (PN) | 102 (0.20) | 90 (0.18) | 0.3353 | 1.16 (0.8-1.6) |
| Symptomatic Irreversible Pulpitis (SIP) | 122 (0.24) | 72 (0.14) | **0.000064** | **1.91 (1.4 - 2.6)** |
| Asymptomatic Irreversible Pulpitis (AIP) | 15 (0.03) | 22 (0.04) | 0.27 | 0.68 (0.3 – 1.3) |
| Reversible Pulpitis (RP) | 4 (0.01) | 3 (0.01) | 0.70 | 1.3 (0.3 – 6) |
| Previously Treated (PT) | 44 (0.09) | 41 (0.08) | 0.73 | 1.08 (0.7 – 1.6) |
| Previously Initiated Therapy (PI) | 13 (0.03) | 8 (0.02) | 0.27 | 1.64 (0.6 – 3.9) |
| All symptomatic (SIP, RP) | 126 (0.25) | 75 (0.15) | **0.000057** | **1.9 (1.3 – 2.6)** |
| All asymptomatic (PN, AIP, PT, PI) | 174 (0.34) | 161 (0.32) | 0.38 | 1.12 (0.8 – 1.4) |
| *Periapical Diagnosis* |  |  |  |  |
| Asymptomatic Apical Periodontitis (AAP) | 20 (0.04) | 15 (0.03) | 0.39 | 1.34 (0.7 – 2.6) |
| Symptomatic Apical Periodontitis (SAP) | 85 (0.17) | 57 (0.11) | **0.01** | **1.6 (1.1 – 2.2)** |
| Acute Apical Abscess (AAA) | 7 (0.01) | 3 (0.01) | 0.20 | 2.3 (0.6 – 9.1) |
| Chronic Apical Abscess (CAA) | 14 (0.03) | 13 (0.03) | 0.84 | 1 (0.5 – 2.3) |
| Normal Apical Tissue (NAT) | 24 (0.05) | 10 (0.02) | **0.01** | **2.4 (1.2 – 5.2)** |
| All symptomatic (SAP, AAA) | 93 (0.18) | 60 (0.12) | **0.003** | **1.7 (1.2 – 2.4)** |
| All asymptomatic (AAP, CAA) | 34 (0.07) | 27 (0.05) | 0.35 | 1.3 (0.7 – 2.1) |
| *Treated Tooth Type* |  |  |  |  |
| Central Incisor | 46 (0.09) | 31 (0.06) | 0.07 | 1.5 (0.9 – 2.4) |
| Lateral Incisor | 27 (0.05) | 39 (0.08) | 0.12 | 0.67 (0.4 – 1.12) |
| Canine | 26 (0.05) | 55 (0.11) | **0.0008** | **2.25 (1.4 – 3.6)** |
| Premolar | 136 (0.27) | 159 (0.32) | 0.1 | 0.8 (0.6 – 1.1) |
| Molar | 264 (0.53) | 216 (0.43) | **0.002** | 1.5 (1.1 – 1.9) |
| All anterior teeth | 99 (0.19) | 125 (0.25) | 0.05 | 0.7 (0.5 – 1) |
| All posterior teeth | 400 (0.8) | 375 (0.75) | 0.06 | 1.3 (0.9 – 1.8) |
| *Jaw* |  |  |  |  |
| Maxilla | 281 (0.56) | 279 (0.56) | 1 | 1 (0.7 – 1.3) |
| Mandible | 218 (0.44) | 221 (0.44) | 0.84 | 0.97 (0.8 – 1.2) |
| *Endodontic Treatment Procedure* |  |  |  |  |
| Root Canal Therapy | 370 (0.74) | 366 (0.73) | 0.77 | 1 (0.8 – 1.4) |
| Endodontic Retreatment | 56 (0.11) | 63 (0.13) | 0.49 | 0.8 (0.6 – 1.2) |
| Pulp Debridement | 30 (0.06) | 41 (0.08) | 0.18 | 0.7 (0.4 – 1.2) |
| Direct/indirect pulp cap; pulpotomy | 34 (0.06) | 21 (0.04) | 0.07 | 1.7 (0.9 – 2.9) |
| Root-end Resection | 10 (0.02) | 15 (0.03) | 0.3 | 0.7 (0.2 – 1.5) |
| Presence of Periapical Lesion | 261 (0.52) | 193 (0.39) | **0.00002** | **1.7 (1.3 – 2.2)** |
| Mean Lesion Size | 2.534 | 2.537 | n.s. | n.s |
| *Healing Outcome* |  |  |  |  |
| Failed/diseased | 24 (0.05) | 53 (0.11) | **0.0006** | **2.35 (1.4 – 3.9)** |
| Healing | 50 (0.1) | 47 (0.09) | 0.7 | 1 (0.7- 1.6) |
| Healed | 73 (0.15) | 89 (0.18) | 0.2 | 0.8 (0.5 -1.1) |

\*Pearson's Chi-square test and/or Fisher's Exact Test. $P \leq 0.05$ indicates statistical significance (in bold). ns, not significant.
\*\* CI, Confidence Interval.
Endo+/DM- and Endo+/DM+, n=500 each.

### Data Analysis

The frequencies of each variable were recorded and compared among the groups. Data analysis was performed using Chi-square and/or Fisher Exact tests, as well as one-way analysis of variance (ANOVA) with posthoc comparisons wherever applicable. All analyses were performed using R statistical software (R Core Team 2018). Multivariate analysis was performed using a generalized linear model with binomial or poisson error distribution to examine the independent and multiplicative effects of age, gender, smoking, periodontal disease, systemic disease and medication considering endodontic treatment, pulpal diagnosis, apical diagnosis, number of treated teeth and healing as response variables. We used the step function in R to assess simplifications of the most complex model. Statistically significant differences were set at P ≤ .05.

## RESULTS

Tables 1 and Supplement Table 1 presents the frequencies obtained for the 24 treatment- and patient-centric variables. Overall, DM+Endo+ presented with less symptomatic pulpal and periapical diagnoses. In particular, the frequency of symptomatic irreversible pulpitis (SIP) and all symptomatic pulpal diagnosis (SIP + RP) was significantly decreased in the DM+ group (P = .00006). Similarly, symptomatic apical periodontitis and all symptomatic periapical diagnosis combined (SAP, AAA) were also significantly decreased in the DM+ group (P<.01) (Figure 1A). The number of endodontically-treated teeth was significantly higher in DM+Endo+ (average 2.6 teeth) in comparison to DM-Endo+ patients (average 1.8 teeth) (P = 2.2 × 10^−16^) (Figure 1B). Endodontic treatment of canines was significantly more frequent in the DM+ group (P=.0008) whereas treatment of molars was more frequent in the DM-group (P=.002) (Figure 1C). The presence of periapical lesions in endodontically-treated teeth was higher in DM-when compared to DM+ (P = 2 × 10^−5^), although no differences were observed for lesion size between groups (P >.05). Diseased/failed treatment outcomes were more frequent in the DM+ group (P = .0006) (Figure 1D).

**Figure 1.**
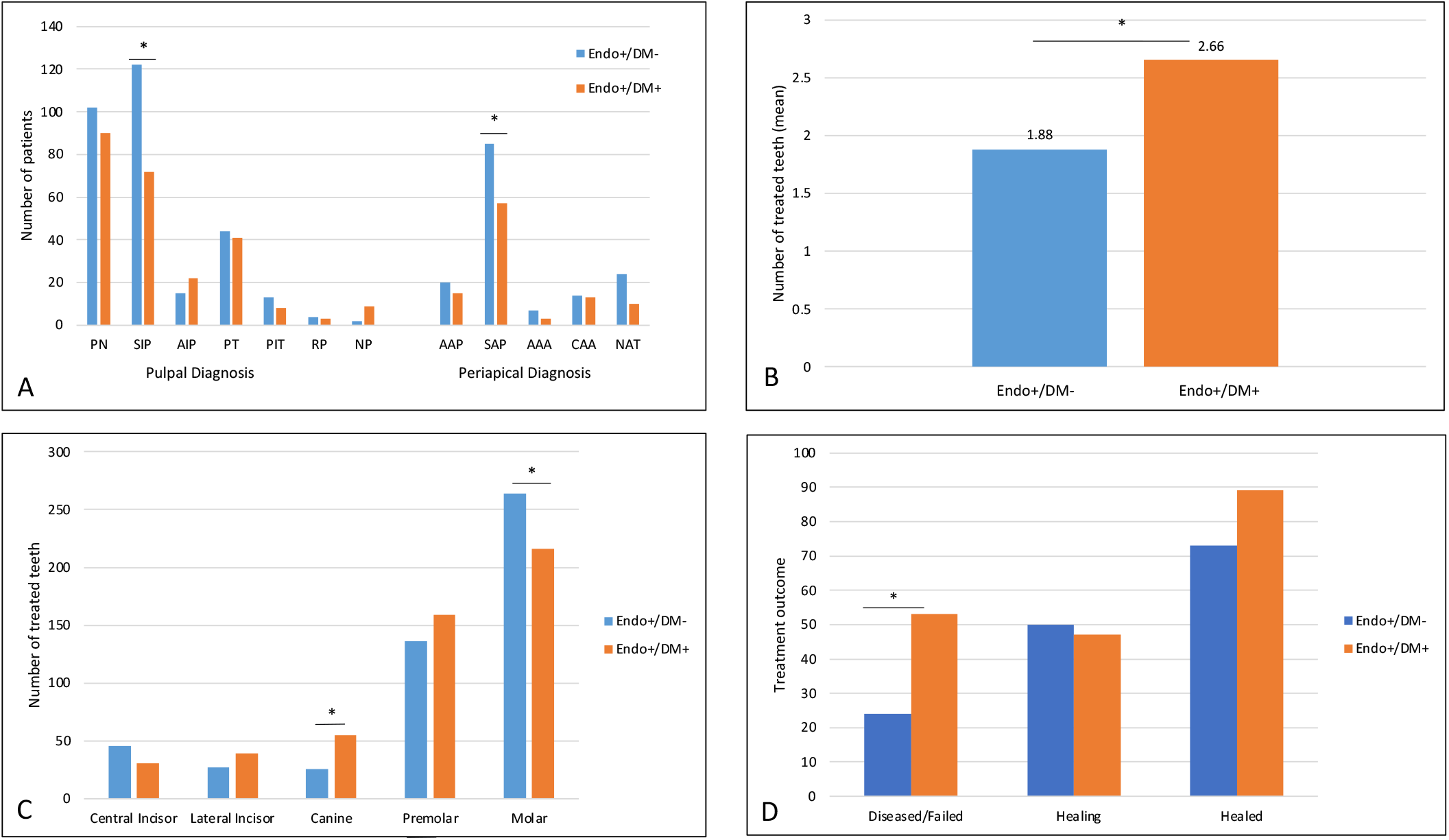
Frequencies of treatment-centric variables evaluated in study groups. **(A)** Pulpal and periapical diagnoses obtained from individual EHR treatment codes. Individuals with DM were less likely to have symptomatic irreversible pulpitis (*P = .000064) or combined symptomatic pulpal diagnoses. Individuals with DM were less likely to have symptomatic apical periodontitis (*P = .01), or combined symptomatic periapical diagnoses (P = .003). **(B)** Average number of endodontically-treated teeth in DM+ and DM-groups. Those DM+ had more treated teeth as compared to DM-individuals (*P = 2.2 × 10^−16^). **(C)** Tooth types frequently treated in DM+ and DM-groups (*P=.0005). **(D)** Healing outcomes in in DM+ and DM-groups. Endo+/DM+ were more likely to have a diseased or failed outcomes (P = .0006). Pulp necrosis (PN), symptomatic irreversible pulpitis (SIP), asymptomatic irreversible pulpitis (AIP), previously treated (PT), previously initiated therapy (PIT), reversible pulpitis (RP), asymptomatic apical periodontitis (AAP), symptomatic apical periodontitis (SAP), acute apical abscess (AAA), chronic apical abscess (CAA), normal apical tissue (NAT).

The presence of periodontal disease was also significantly increased in the Endo+DM+ group (P = .01), and represented mostly cases of localized chronic periodontitis (P = .006) (Figure 2A). The presence of an additional systemic disease was significantly higher in the DM+ group, regardless of endodontic treatment status (P = .005) (Figure 2B). Similarly, hypertension (HTN) and obesity were the most frequently reported disease among groups, and significantly overrepresented in DM+ groups regardless of endodontic treatment (P = 1 × 10^−5^). Coronary artery disease (CAD), congestive heart failure (CHF), hyperlipidemia, and hypothyroidism, were also significantly higher in DM+ patients (P > .05) (Figure 2C). Smoking was lower in the Endo+DM+ group (P = 0.04). No significant differences were found when comparing the frequencies of family history of diabetes, or number of medications taken (P ≥ .05). No significant differences were found comparing age or gender between groups (P ≥ .05) (Supplement Table 1).

**Figure 2.**
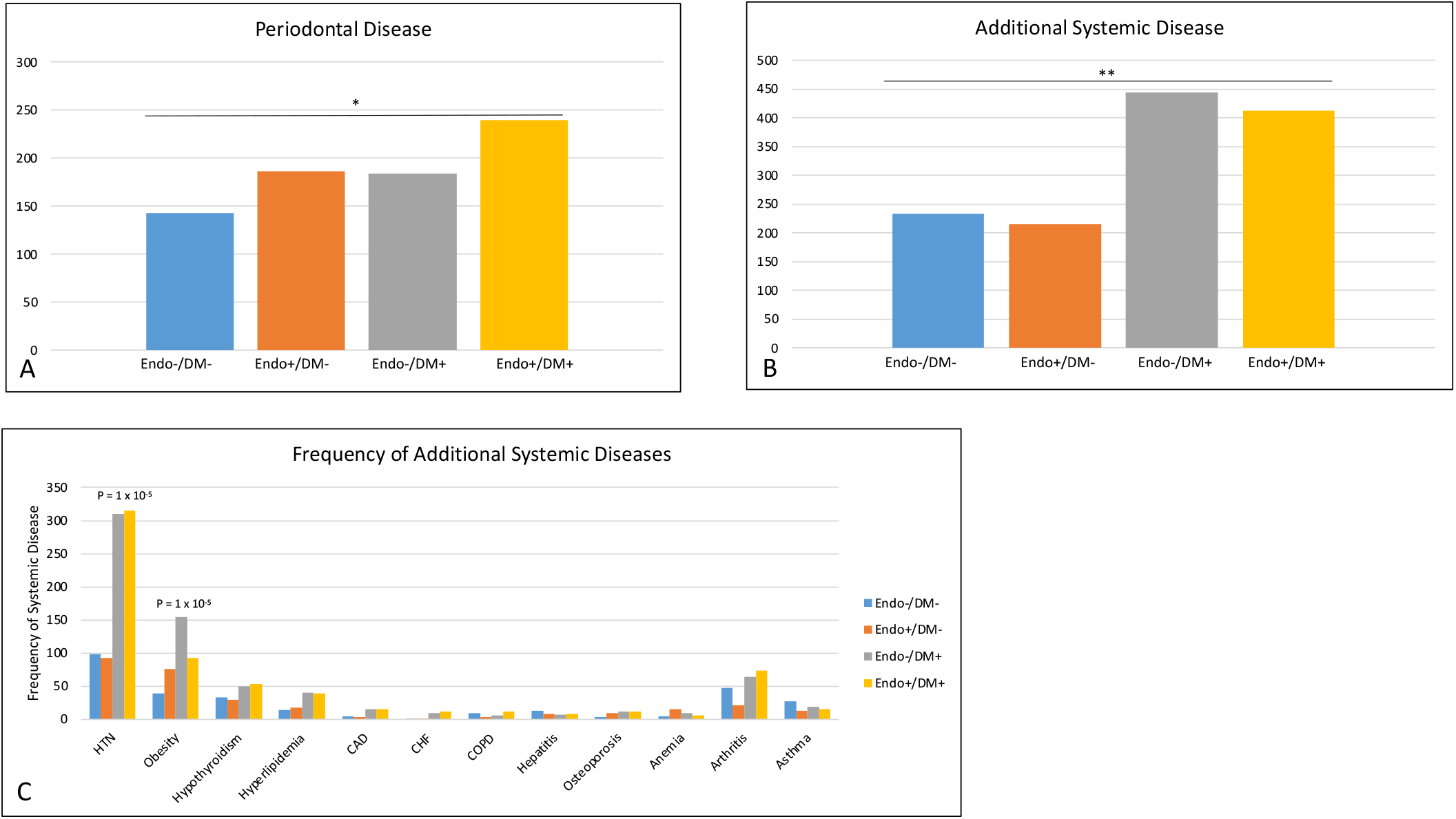
Frequency of additional diseases in the study groups. **(A)** Frequency of periodontal disease among groups (*P=.0123). **(B)** Frequency of additional systemic diseases in study groups. DM+ groups have more additional systemic diseases (*P = .005). **(C)** Frequency of individual systemic diseases in DM+ and DM-groups. Hypertension (HTN), obesity, hypothyroidism, coronary artery disease, congestive heart failure, were significantly different among groups (P<.05).

Multivariate analysis results

## DISCUSSION

In this study, we evaluated the potential relationship between DM and endodontic pathosis through a retrospective analysis of 2,000 electronic health records. For each group, we compared information on 24 treatment- and patient-centric variables, to determine the potential association between DM and endodontic pathosis presentations and treatment outcomes. Our results continue to support that DM contributes to increased endodontic disease and highlight the complex relationships between oral and systemic diseases.

The number of endodontically-treated teeth was significantly higher in diabetic individuals (2.66 vs. 1.88), with canines being treated more often compared with non-diabetics. While the frequency of endodontic treatment in diabetics is not commonly reported, our results corroborate previous findings in which 70% of a diabetic population had one or more endodontically-treated teeth, whereas only 50% of those without DM had endodontic treatment.^27^ In contrast, similar numbers of endodontically-treated teeth in individuals with and without DM have been reported.^28^

Our findings also showed significant differences for pulpal and periapical diagnoses between groups, with a significantly lower frequency of symptomatic pulpal and periapical diagnoses in DM+ patients. A reduced sensibility in DM+ patients may impact the results of endodontic diagnostic testing and in turn skew treatment decision and modality. In agreement with our findings, asymptomatic pulpal/periapical infections were previously reported as more frequent in DM patients.^6,29^ In contrast, an increasing trend towards symptomatic apical periodontitis among individuals with type I DM and DM has also been described.^30^ By distinguishing between type I and type II DM, Fouad was able to determine that type I DM only shows a trend toward increased symptomatic AP.^30^ Of note, our study did not differentiate between type I DM, type II DM, or pre-DM, although we expect a majority of type II DM in our sample, in line with observed distribution in the general population.

With regards to endodontic procedure types, no differences were observed between DM+ and DM-groups. Albeit nonsignificant, endodontic retreatment and root-end resection were most commonly noted for DM+ patients. Decreased endodontic treatment success and delayed healing rates have been reported in DM+ patients, particularly in the presence of apical periodontitis.^29–33^ Further, a 1.29 hazard ratio of extraction of endodontically-treated teeth has been reported for DM+ patients, and was more evident in the presence of an additional systemic disease.^34^

Previous evidence consistently suggest an increased prevalence of AP and larger periapical lesion sizes in DM patients.^16,17,27–29^ In rat models of AP, the presence of apical lesions and higher morbidity/mortality rates have often been associated with DM.^35,36^ Interestingly, our results do not corroborate these previous findings. Such discrepancies may be due to differences in study designs therefore direct comparisons are not possible. In the present study, we assessed the presence of active AP lesions in endodontically-treated teeth and found that the number of active periapical lesions was higher in the DM-group than in the DM+, meanwhile lesion sizes were similar among groups. We speculate that this may be due to a more stringent follow-up regimen adopted by diabetic patients, and the proactive treatment approaches in a dental school setting. Neumann et. al. (2017) found that while typically only 50% of diabetic patients undergo regular dental examinations, they found that 83% of those underwent a dental procedure.^37^

Similar to most studies, the timing of endodontic treatment relative to the onset of endodontic disease and/or control of DM and other systemic diseases were not able to be evaluated in this study.^30^ A recent review by Nagendrababu et al. (2020) found DM to be a preoperative prognostic factor in endodontic outcomes, but emphasized the need to consider additional variables such as systemic medication use.^5^ Commonly used medications among diabetics such as metformin and statins are known to improve AP and tissue repair, therefore knowledge of past and current medications taken by DM patients should be considered when evaluating AP lesion sizes and improved treatment outcomes.^5,38,39^ In this context, our findings showed a significant trend towards failed and/or diseased outcomes in DM+ individuals. Although information on healing outcome was not readily available for all patients, our findings are in line with previous reports. ^30–33^ Fouad et al. (2003) showed an increased risk of flare-up and reduced healing outcomes if a periapical lesion was present in DM patients.^30^ A potential caveat in this study was the use of periapical radiographs to assess the presence of periapical lesions. Low et al. (2008) reported that periapical radiographs were 34% less accurate to detect periapical lesions compared to cone beam computed tomography (CBCT). ^40^ Patel et al. (2007) also demonstrated that only 20% of periapical lesions (sized 2mm or less) of mandibular first molars were identified by periapical radiographs.^40,41^ While the use of CBCT would be ideal for a more accurate representation of actual lesion sizes, this was not possible due to the retrospective nature of this study and the limited number of CBCT images available during the period of data collection.

In an effort to determine the additive role of other diseases/conditions in DM and endodontic pathologies, we compared the frequencies of periodontal disease and additional systemic conditions among the groups. Our results showed a significant association between periodontal disease in the Endo+/DM+ group. A relationship between DM and periodontal disease is well established in the literature, and our results are in agreement.^12,25,42–46^ Interestingly, DM+ patients were noted to have at least one additional systemic disease in comparison to DM-patients. In line with these findings, the use of certain medications was also more frequent among DM+ patients. As highlighted above, knowledge of past and current systemic medication use by endodontic patients is paramount as certain medications are known to influence endodontic outcomes and AP.^5^ Our findings suggest a significantly higher incidence of cardiovascular disease (CVD) among DM+ patients, particularly coronary artery disease (CAD), congestive heart failure (CHF), hyperlipidemia, and hypertension (HTN). The use of lipid lowering agents, antihypertensives, and anticoagulant drugs was also higher among DM+ patients. CVDs have been described in association with endodontic disease in numerous studies,^47–50^ and while the exact mechanisms underlying the link between these conditions are poorly understood, these findings highlight the complexity of potential oral health-systemic health relationships.

We also tested for the association with smoking status as smoking aggravates DM-related complications; high levels of nicotine can decrease insulin effectiveness thereby further dysregulating the process of glycemic control.^51^ Only a trend for association was found between smoking, and driven by the lower frequency of smokers in the Endo+/DM+ group. Of note, available data on smoking was categorical and did not allow for evaluation of smoking over the course of an individual’s life. It has been previously shown that the incidence of endodontic treatment failures is higher among smokers (independent of DM status).^32^ Indeed, while our study also found a higher trend towards failed treatments in DM+ patients, a relationship between smoking, DM, and healed/failed outcome could not be established.

The limitations of this study include the retrospective nature using patient self-reported history, and the potential sources of bias introduced in data entry by the care providers. It may also be possible that the criteria utilized in the study would not identify patients who are diabetic but not aware of it. Nevertheless, among the strengths of this study, we highlight the large sample size and the use of standardized diagnostic and procedure codes used for data retrieval. To our knowledge, this is the first study to systematically evaluate treatment- and patient-centric variables to better understand the potential effects of DM on endodontic pathologies.

## CONCLUSIONS

Within the limitations of this study, DM was found to increase the frequency of endodontic treatment, meanwhile reducing the frequency of symptomatic pulpal and periapical diagnoses. Future prospective studies should consider assessing the type of DM, duration and control of disease, and the timing of endodontic treatment to further establish the effects of DM on AP.

## Data Availability

All data produced in the present work are contained in the manuscript

## ACKNOWLEDGEMENTS

Thanks to Dr. Nat Holland, PhD, for help with statistical analyses.

## CONFLICT OF INTEREST

The authors deny any conflicts of interest to this study.

## Notes

### Competing Interest Statement

The authors have declared no competing interest.

### Funding Statement

This study did not receive any funding

### Author Declarations

Ethics committee/IRB of the UTHealth Committee for Protection of Human Subjects gave ethical approval for this work(HSC-DB-17-0565).

