## Supplementary material for "Association Between Diabetes Mellitus and Endodontic Pathosis"

**Supplement Table 1.** Frequencies of patient/disease-specific variables between groups.

| Variable | Endo-/DM- | Endo+/DM- | Endo-/DM+ | Endo+/DM+ | P-value* |
|---|---|---|---|---|---|
| *Gender* | | | | | 0.4647 |
| Female | 308 | 311 | 283 | 270 | |
| Male | 186 | 186 | 212 | 229 | |
| Unknown (not reported) | 6 | 3 | 5 | 1 | |
| Average age, years (min-max) | 49.5 (29-101) | 50 (29-93) | 66 (30-98) | 65 (29-98) | 0.6916 |
| Presence of Additional Systemic Disease | 233 | 216 | 445 | 413 | **0.005** |
| *Type of Systemic Disease* | | | | | |
| Adrenal Gland Disorder | 0 | 0 | 1 | 0 | ns |
| Anemia | 5 | 15 | 10 | 6 | ns |
| Angina | 0 | 1 | 0 | 0 | ns |
| Arthritis | 47 | 21 | 64 | 74 | ns |
| Atrial Fibrillation | 1 | 4 | 3 | 0 | ns |
| Asthma | 27 | 13 | 19 | 15 | ns |
| Blood Disorders | 2 | 2 | 0 | 1 | ns |
| Benign Prostate Hyperplasia | 0 | 0 | 2 | 0 | ns |
| Bradycardia | 0 | 1 | 0 | 0 | ns |
| Coronary Artery Disease | 5 | 4 | 15 | 15 | **0.009** |
| Cardiac Arrhythmia | 2 | 2 | 4 | 0 | ns |
| Cerebral Palsy | 0 | 0 | 1 | 0 | ns |
| Congestive Heart Failure | 1 | 1 | 10 | 12 | **0.0006** |
| Cirrhosis | 1 | 3 | 1 | 1 | ns |
| Chronic obstructive pulmonary disease | 10 | 3 | 6 | 12 | ns |
| Dementia | 0 | 0 | 0 | 1 | ns |
| Deep Vein Thrombosis | 2 | 1 | 0 | 0 | ns |
| Fibromyalgia | 0 | 1 | 0 | 1 | ns |
| Gout | 0 | 0 | 0 | 6 | ns |
| Goiter | 0 | 0 | 1 | 0 | ns |
| Graves | 0 | 1 | 0 | 1 | ns |
| Hepatitis | 13 | 8 | 7 | 8 | ns |
| Hypertension | 99 | 93 | 310 | 315 | **$1 \times 10^{-5}$** |
| Huntington's | 0 | 0 | 1 | 0 | ns |
| Hyperlipidemia | 14 | 18 | 40 | 39 | **$9 \times 10^{-5}$** |
| Hyperthyroidism | 4 | 2 | 1 | 3 | ns |
| Hypothyroidism | 33 | 30 | 50 | 53 | **0.0132** |
| Irritable Bowel Disease | 5 | 0 | 1 | 2 | ns |
| Kidney Failure | 6 | 0 | 26 | 5 | ns |
| Lichen Planus | 1 | 0 | 1 | 0 | ns |
| Lupus | 0 | 1 | 1 | 2 | ns |
| Myocardial Infarction | 2 | 3 | 8 | 4 | ns |
| Multiple Sclerosis | 1 | 1 | 0 | 0 | ns |
| Murmur | 3 | 1 | 1 | 2 | ns |
| Mitral Valve Prolapse | 0 | 0 | 0 | 4 | ns |
| Osteoporosis | 4 | 9 | 12 | 12 | ns |
| Obesity | 39 | 76 | 154 | 93 | **$1 \times 10^{-5}$** |
| Palpitations | 6 | 4 | 2 | 0 | ns |
| Parkinson's | 0 | 0 | 1 | 0 | ns |
| Polycystic Ovarian Syndrome | 1 | 1 | 0 | 0 | ns |

| | | | | | |
|---|---|---|---|---|---|
| Sjogren's Syndrome | 0 | 2 | 0 | 1 | ns |
| Periodontal Disease | 143 | 186 | 184 | 240 | **0.0123** |
| *Periodontal Classification* | | | | | **0.0055** |
| Healthy Periodontium | 7 | 13 | 5 | 10 | |
| Gingivitis | 55 | 124 | 40 | 76 | |
| Generalized Chronic Periodontitis | 71 | 103 | 124 | 121 | |
| Localized Chronic Periodontitis | 24 | 36 | 31 | 62 | |
| Family History of Diabetes | 180 | 167 | 328 | 325 | 0.8311 |
| Smoking | 93 | 99 | 93 | 67 | **0.0438** |
| *Medication Intake* | | | | | |
| +1 Medication | 72 | 94 | 211 | 265 | 0.5868 |
| +2 Medication | 38 | 51 | 189 | 239 | 0.7344 |
| +3 Medication | 23 | 36 | 163 | 198 | 0.5809 |
| *Medication Type* | | | | | |
| Allergy & Autoimmune | 12 | 10 | 18 | 28 | **0.0076** |
| ADHD | 2 | 1 | 3 | 8 | **0.0394** |
| Anticoagulants & Antiplatelets | 2 | 8 | 16 | 5 | **0.0026** |
| Antidepressants | 14 | 20 | 22 | 46 | **2 x 10-$^{5}$** |
| Antidiabetics | 0 | 4 | 196 | 286 | ns |
| Antifungals | 1 | 1 | 4 | 4 | ns |
| Antigout | 0 | 2 | 5 | 9 | ns |
| Antihypertensives | 41 | 66 | 231 | 272 | **1 x $10^{-5}$** |
| Antipsychotic | 3 | 0 | 1 | 10 | ns |
| Antitumor | 0 | 0 | 3 | 2 | ns |
| Antivirals | 3 | 1 | 3 | 2 | ns |
| Antacids, Nausea | 5 | 12 | 35 | 35 | **1 x $10^{-5}$** |
| Aspirin | 8 | 10 | 35 | 46 | **1 x $10^{-5}$** |
| Antianxiety, Sedatives, Sleep-aids | 2 | 6 | 8 | 19 | **0.0004** |
| Bladder Drugs | 2 | 0 | 3 | 3 | ns |
| Cognition Drugs | 1 | 0 | 3 | 2 | ns |
| Diuretics | 2 | 6 | 22 | 17 | **5 x $10^{-5}$** |
| Eye Drops | 3 | 3 | 1 | 14 | **0.0002** |
| Gastrointestinal Tract | 0 | 2 | 8 | 4 | ns |
| Heart Regulators | 3 | 1 | 6 | 6 | ns |
| Hormones | 5 | 6 | 4 | 6 | ns |
| Immunosuppressants | 0 | 3 | 3 | 0 | ns |
| Insulin | 0 | 0 | 43 | 49 | ns |
| Lipid-Lowering Agents | 19 | 22 | 112 | 128 | **1 x $10^{-5}$** |
| Migraine & Seizure | 3 | 1 | 4 | 8 | ns |
| Muscle Relaxers | 0 | 3 | 6 | 8 | ns |
| Nonsteroidal anti-inflammatory | 4 | 4 | 17 | 10 | **0.004** |
| Oral Solutions (i.e. Prevident) | 0 | 1 | 1 | 1 | ns |
| Prostate Drugs | 3 | 2 | 15 | 20 | **2 x $10^{-5}$** |
| Opioids | 5 | 5 | 38 | 32 | **1 x $10^{-5}$** |
| Stimulants | 0 | 0 | 1 | 0 | ns |
| Thyroid | 12 | 16 | 28 | 42 | **3 x $10^{-5}$** |
| Vitamin, Mineral, Regulating Drugs | 8 | 7 | 27 | 6 | **1 x $10^{-5}$** |
| Weight Regulating Drugs | 1 | 2 | 2 | 0 | ns |

*Pearson's Chi-squared test, analysis of variance, and Fisher's Exact Test. $P \leq 0.05$ indicates statistical significance.

**Supplement Table 2.** Results of multivariate analyses.

| Pr(>\|z\|) | Estimate | Std. error | Z value | Intercept |
|---|---|---|---|---|
| dmyes | | | | |
| age | | | | |
| genderM | | | | |
| smokeYes | | | | |
| onemedYEs | | | | |
| perioYes | | | | |
| sysdzYes | | | | |
| dmYes:age | | | | |
| dmYes:smokeYes | | | | |
| age:smokeYes | | | | |
| genderM:smokeYes | | | | |
| dmYes:onemedYes | | | | |
| age:onemedYes | | | | |
| genderM:onemedYes | | | | |
| smokeYes:onemedYes | | | | |
| dmYes:perioYes | | | | |
| age:perioYes | | | | |
| genderM:perioYes | | | | |
| smokeYes:perioYes | | | | |
| onemedYes:perioYes | | | | |
| dmYes:sysdzYes | | | | |
| age:sysdzYes | | | | |
